# Whole Exome and Transcriptome Sequencing of Stage-Matched, Outcome-Differentiated Cutaneous Squamous Cell Carcinoma Identifies Gene Expression Patterns Associated with Metastasis and Poor Outcomes

**DOI:** 10.1101/2024.02.05.24302298

**Authors:** Shams Nassir, Miranda Yousif, Xing Li, Kevin Severson, Alysia Hughes, Jacob Kechter, Angelina Hwang, Blake Boudreaux, Puneet Bhullar, Nan Zhang, Duke Butterfield, Tao Ma, Ewoma Ogbaudu, Collin M Costello, Steven Nelson, David J DiCaudo, Aleksandar Sekulic, Christian Baum, Mark Pittelkow, Aaron R Mangold

**Author notes:** – Corresponding Author. – Equal Contribution (Shams Nassir, Miranda Yousif, Aaron R Mangold).

## Abstract

Cutaneous squamous cell carcinoma (cSCC) is one of the most common cancers in humans and kills as many people annually as melanoma. The mutational and transcriptional landscape of cSCC has identified driver mutations associated with disease progression as well as key pathway activation in the progression of pre-cancerous lesions. The understanding of the transcriptional changes with respect to high-risk clinical/histopathological features and outcome is poor. Here, we examine stage-matched, outcome-differentiated cSCC and associated clinicopathologic risk factors using whole exome and transcriptome sequencing on matched samples. Exome analysis identified key driver mutations including *TP53*, *CDKN2A*, *NOTCH1*, *SHC4*, *MIIP*, *CNOT1*, *C17orf66*, *LPHN22*, and *TTC16* and pathway enrichment of driver mutations in replicative senescence, cellular response to UV, cell-cell adhesion, and cell cycle. Transcriptomic analysis identified pathway enrichment of immune signaling/inflammation, cell-cycle pathways, extracellular matrix function, and chromatin function. Our integrative analysis identified 183 critical genes in carcinogenesis and were used to develop a gene expression panel (GEP) model for cSCC. Three outcome-related gene clusters included those involved in keratinization, cell division, and metabolism. We found 16 genes were predictive of metastasis (Risk score ≥ 9 Met & Risk score < 9 NoMet). The Risk score has an AUC of 97.1% (95% CI: 93.5% - 100%), sensitivity 95.5%, specificity 85.7%, and overall accuracy of 90%. Eleven genes were chosen to generate the risk score for Overall Survival (OS). The Harrell’s C-statistic to predict OS is 80.8%. With each risk score increase, the risk of death increases by 2.47 (HR: 2.47, 95% CI: 1.64-3.74; p<0.001) after adjusting for age, immunosuppressant use, and metastasis status.

## Introduction

Non-melanoma skin cancers (NMSC) are the most common cancer in humans, and the fifth most costly [1–3]. cSCC reportedly metastasizes in up to 5% of patients and kills as many people annually as melanoma [4]. Current staging systems, American Joint Committee on Cancer (AJCC)-8 and Brigham Women’s Hospital (BWH), use clinical and histopathological risk factors to assess risk of recurrence or metastasis [5–7]. Both staging systems have defined intermediate to high-risk tumors as T2 or higher. The BWH further refines T2 lesion classification into T2a and T2b based on the number of clinical and histopathological risk factors present. T2a tumors (18.3% of all cSCC) are identified as intermediate risk, with a local recurrence rate of 5%, lymph node involvement in up to 7%, and disease-specific death in 1% [8]. Although T2a tumors are less likely to have poor outcomes, there are a subset of tumors within this group that have a high risk for local recurrence and nodal metastasis. For example, T2a tumors that invade the subcutaneous fat can spread to lymph nodes in up to 22% of cases [8]. Thus, further refinement of the T2a classification is needed. T2b tumors (4.7% of all cSCC) are considered high risk with a local recurrence rate of 21%, lymphatic metastasis in up to 30%, and disease-specific death in 10% [8]. Patients with T2b tumors and T3 have been defined as a group that may benefit from pre-operative imaging, a sentinel lymph node biopsy, and post-operative radiation [8, 9]. Progress in understanding altered genes in cancer has led to the identification of improved prognostic biomarkers [10]. Whole exome sequencing (WES) of aggressive cSCC (characterized by frequent recurrences, increased risk of metastasis, and high disease-related mortality) identified *TP53, CDKN2A, NOTCH1,2,* and *AJUBA* as candidate driver genes [11–15]. WES of well- and moderately-differentiated cSCC also identified phenotype-specific (e.g., high-risk histological features) candidate driver genes and biological functions [11, 14]. WES of lymph node metastases of cSCC identified *CDKN2A, TP53, and NOTCH1* as candidate driver genes, and showed that dysregulation of RAS/TRK/PI3K, cell cycle, and cellular differentiation pathways is present in the metastatic setting [13].

Gene expression arrays have demonstrated transcriptomic differences between cSCC and actinic keratoses (AK) as well as between different histological phenotypes of cSCC [12]. The *MAPK* pathway was found to play a role in the transition from pre-cancer to malignancy, and unsupervised hierarchal clustering revealed that well-differentiated tumors cluster separately from moderately and poorly differentiated tumors [12, 16]. However, understanding of the transcriptional changes with respect to high-risk clinical/histopathological features and outcome is poor. Multi-gene prognostic modeling in high-risk cSCC has demonstrated promise in early reports [17, 18]. To date, there have been no prognostic models integrating WES and whole transcriptomic sequencing on matched samples. Herein, we perform integrated genomics and transcriptomics on a homogenous group of stage-matched cSCC tumors with known outcomes (metastasis versus no-metastasis) and present the first prognostic model developed using multi-omic data.

## Methods

### Case identification and Histopathological Review

This study was approved by the Mayo Clinic Institutional Review Board IRB 21-012833. A total of 61 cases of cSCC were identified from our retrospective cSCC database comprised of patients receiving care at Mayo Clinic (Jacksonville, Florida; Rochester, Minnesota; and Scottsdale, Arizona). These were identified through an enterprise-wide search for cases with a histopathologic diagnosis of cSCC from pathology reports of archived specimens. Tissue samples were selected based on clinical outcomes (local recurrence, regional metastasis [including nodal and in-transit metastasis], and distant metastasis), histopathologic features, and tissue availability. Tissue samples were selected consecutively between January 1, 1999, and February 2, 2022. All tissues identified at inclusion were BWH stage T2a and T2b prior to histopathological review. All cases underwent histopathologic rereview by board-certified dermatopathologists to confirm pathological staging (D.J.D. and S.A.N.) and had at least 18-months of clinical follow-up data. Clinical characteristics were collected from the electronic medical record, including age, sex, immunosuppression status, and tumor stage (BWH and AJCC seventh edition). Reviewers were blinded to outcome.

### Whole Exome Sequencing and Analysis

#### Tissue identification, preparation, and exome sequencing

We performed whole exome sequencing (WES) on 61 primary tumors (32 metastatic and 29 non-metastatic) and 59 normal controls from a cohort of 59 patients (two patients with multiple primary tumors). Primary tumor and normal control tissues were macrodissected from serial unstained slides as guided by a board-certified dermatologist. DNA was isolated from formalin-fixed paraffin-embedded (FFPE) tissues using the GeneRead FFPE kit following manufacturer recommendations (Qiagen). Paired-end libraries were prepared with as low as 10 ng of FFPE DNA using the SureSelect XT Low Input Reagent Kit (Agilent, Santa Clara, CA). Briefly, adaptor-ligated DNA was amplified with the SureSelect Pre-Capture forward and specific index reverse primers for 12 cycles. The concentration and size distribution of the amplified libraries were determined using an Agilent Bioanalyzer DNA 1000 chip or Advance Fragment Analyzer and Qubit fluorometry (Invitrogen, Carlsbad, CA). Whole exon capture was then carried out using 750 ng of the prepped library, following the protocol for Agilent’s SureSelect Human All Exon v5 + UTRs 75 MB kit. The purified capture products were amplified using the SureSelect Post-Capture Indexing forward and Index PCR reverse primers for 12 cycles. The concentration and size distribution of the captured libraries were determined using Qubit fluorometry and the Agilent Bioanalyzer DNA 1000 chip. Libraries were sequenced at an average coverage of approximately 80x following Illumina’s standard protocol using the Illumina cBot and HiSeq 3000/4000 PE Cluster Kit. The flow cells were sequenced as 150 × 2 paired end reads on an Illumina HiSeq 4000 using the HiSeq Control Software HD 3.4.0.38 collection software. Base-calling was performed using Illumina’s RTA version 2.7.7.

#### Alignment and Somatic variant calling

Genome_GPS v5.0.3 (formerly known as TREAT) was used as a comprehensive secondary analysis pipeline for DNA sequencing data [19]. FASTQ files were aligned to the hg38 reference genome using bwa-mem (v0.7.10) with default settings. Realignment was performed using GATK (v3.4–46) [20]. To identify somatic mutations, the workflow employed a combination of Mutect2 and Strelka2 [21, 22]. Identified variants were annotated using BioR framework, which included functional features, impact prediction, and clinical significance assessments using databases such as Clinical Annotation of Variants, ClinVar, Human Gene Mutation Database, Mayo Biobank, and Exome Aggregation Consortium population frequencies [23].

#### Somatic variant filtering

Raw variants were filtered based on the following criteria: minimum alternate allele fraction of 10%, minimum number of reads supporting the alternate allele of 2, and a minimum read depth of 10. Additionally, variants located in repetitive element regions defined by RepeatMasker (v4.1.2) or the Simple Repeat track in the UCSC Genome Browser were excluded. OxoG artifacts were removed using the Picard tools CollectOxoGMetrics and FilterByOrientationBias (v2.21.6). Common variants were further eliminated based on their minor allele frequency (>= 0.01) as recorded in the 1000 Genomes Project.

#### Driver genes identification

To identify coding driver genes, three discovery algorithms were employed: MutSigCV (v1.41), OncodriveFML (v2.4.0), and OncodriveCLUSTL (v1.1.1) [24–26]. MutSigCV was used to detect driver genes based on recurrence, OncodriveFML was used to identify drivers enriched for mutations with high functional impact using whole exome mutation frequencies to model the background mutation rate, and OncodriveCLUSTL was used to detect drivers containing clusters of mutations. The default parameter settings were used for all three algorithms. A p-value was generated for each gene by each algorithm, and genes were considered as drivers when p < 0.05 for at least two out of the three algorithms.

### Whole Transcriptome Sequencing and Analysis

#### Tissue preparation and RNA sequencing

We performed whole transcriptome sequencing on a total of 53 primary tumors including 22 metastatic and 31 non-metastatic lesions from 50 patients (3 patients had multiple primary tumors). Twenty-two tumor-adjacent normal controls were sequenced and utilized for differential gene expression analysis. Primary tumor and normal control tissues were macrodissected from serial unstained slides as guided by a board-certified dermatologist. RNA was isolated from FFPE tissue using the RNeasy FFPE kit (Qiagen) following manufacturer recommendations. Total RNA concentration and quality were determined using Qubit fluorometry (Invitrogen, Carlsbad, CA) and the Agilent Fragment Analyzer (Santa Clara, CA). Samples with a DV200 of 30% or better proceeded to library prep. Using the Illumina TruSeq® RNA Exome Library Prep kit (San Diego, CA), libraries were prepared according to the manufacturer’s instructions using up to 500 ng of FFPE RNA. The concentration and purity of cDNA libraries were checked using the TapeStation D1000 (Agilent, Santa Clara, CA). Coding regions of the transcriptome were captured by pooling four of the cDNA libraries at 200 ng each. The concentration and size distribution of the completed libraries were determined using an Agilent Bioanalyzer DNA 1000 chip (Santa Clara, CA) and Qubit fluorometry (Invitrogen, Carlsbad, CA). Libraries were sequenced at 8 samples per lane following Illumina’s standard protocol using the Illumina cBot and HiSeq 3000/4000 PE Cluster Kit. The flow cells were sequenced as 100 × 2 paired end reads on an Illumina HiSeq 4000 using the HiSeq Control Software HD 3.4.0.38 collection software. Base-calling was performed using Illumina’s RTA version 2.7.7.

All samples were sequenced at the Mayo Clinic Medical Genome Facility Sequencing Core by Illumina HiSeq 4000 with paired end 101-base pair (bp) read length. Approximately 60 million reads per sample were generated. MAP-Rseq v3 was used to analyze RNA-Sequencing data [27]. The aligning and mapping of reads were performed using TopHat2 against the human reference genome (hg38). Gene counts were generated by FeatureCounts using the gene definitions files from Ensembl. RseqQC was used to create a variety of quality control plots to ensure the results from each sample were reliable for the downstream differential expression analysis.

The R software package DESeq2 was used for differential gene expression analysis [28]. Differentially expressed genes (DEGs) between SCCs (n=53) and controls (n=22) are identified by adjusted p-values <0.0001 and log 2-fold change >1 (up-regulated) or <-1 (down-regulated). Pathway enrichment analysis was performed using the Reactome pathway database on up- and down-regulated genes. DEGs in the top enriched pathways with FDR-corrected Benjamin p value <0.05 were extracted for gene-gene interaction network analysis using STRING gene/protein interaction database. We used StringDB to identify regulatory hub genes (those with the highest number of known gene interactions) within these pathways [29]. The hub genes were identified by the number of interactions those genes have with the others. We selected top 50 up-regulated and down-regulated hub genes for establishing SCC molecular panel. Similarly, DEGs between Met and NoMet groups are identified by FDR-adjusted p values < 0.05 and log 2-fold change >1 (up-regulated) or less than <-1 (down-regulated). Fourteen of them also carry mutations identified by exome sequencing data and were included in the SCC molecular panel.

#### Integration of RNA and DNA for gene expression panel (GEP)

We compiled a list of 183 genes for a cSCC gene expression panel (GEP) through multi-omic data integration, including high-interest genetic mutations identified through exome sequencing as well as results identified through various transcriptomics approaches as outlined in previous sections. To identify functionally relevant mutated genes, the list of somatic mutations was further filtered through the selection of mutations, including driver mutations by two calling systems, high Clinical Annotation of VAriants (CAVA) score with a threshold cutoff of 10 mutations, top 20 mutated genes [initially using a nested cohort of 45 cases for optimization (Data not shown) and 61 cases in final derivation], top 50 minor allelic frequency and gene expression changes of log 2-fold change > 1 or < −1 [30]. Transcriptomic data was selected based on the top 50 up- and down-regulated hub gene selection of enriched pathways identified above as well as the key differential genes between metastatic and non-metastatic tumors.

### Data normalization and modeling

#### Expression value tiering for modeling analysis

The gene expression data of 183 genes were pulled from the raw data of standard secondary analysis pipeline. The expression values were then sorted by log 2-scale. We developed expression tiers by comparing each gene’s expression value in each tumor sample to its respective mean expression in the control group. Genes with expression values at least one standard deviation greater than the control group mean were assigned a value of 1; those with expression values at least one standard deviation less than the control group mean were assigned a value of −1. The remaining genes were assigned a value of 0. In summary, the genes are categorized as follows:D1 (expr > mean + n*sd), −1 (expr < mean - n*sd), and otherwise 0,Dwhere n is either 1 or 0.D

#### Gene expression profiling of selected genes for survival and metastasis

Partitioning Around Medoids (PAM) method was used to cluster the genes to investigate the correlations between them [31]. Gap statistic was used as the criteria to choose the optimal number of clusters. Pearson correlation coefficients using the genes’ normalized raw expression data were calculated and presented in a heat map. To choose the genes that are potentially predictive of metastasis, logistic least absolute shrinkage and selection operator (LASSO) regression was used. 100 cross-validation logistic LASSO regression was used to screen the genes that can be associated with metastasis. The genes that were chosen 50 times or more were further considered [32]. For the genes that were chosen by the LASSO regression, univariate analysis with metastasis status was further investigated using Fisher’s exact test. The genes that were significant from Fisher’s exact test and had up-regulated or down-regulated associations with metastasis were chosen to generate the risk score for metastasis. For the generated risk score, receiver operating characteristic curve (ROC) analysis was conducted and Youden index was used to choose the optimal cut-off point to predicting metastasis. The area under the ROC curve (AUC) was calculated. The sensitivity, specificity and overall accuracy of the optimal cut-off point for predicting metastasis were also calculated.

Regularized Cox regression was used to choose the genes that are potentially associated with overall survival [33]. A similar modeling strategy was applied – 100 cross-validation regularized Cox regression was used to choose the genes associated with overall survival; among the 100 cross-validation models, the genes selected 50 times or more were further considered. For the genes that were chosen by the LASSO Cox regression, log-rank test and Kaplan-Meier curves were used to investigate their association with overall survival. DEGs associated with worse overall survival were chosen to generate risk scores for overall survival.

The risk scores were combined into low-risk, medium-risk, and high-risk groups based on their Kaplan-Meier curves. The median survival time for each risk group was further estimated. Harrell’s C-statistics for the risk score predicting overall survival was calculated. Multivariable Cox regression adjusting for age, immunosuppressant use, and metastasis status was used to investigate the association between the risk score and overall survival.

## Results

A total of 61 cases were selected for exome sequencing (Table 1). Fifty patients were selected for transcriptomic sequencing; patient demographics are shown in Supplemental Table 1. Detailed RNA-seq sample group information is shown in Supplemental Table 2.

**Table 1:** Patient demographics-The 61 DNA samples are from 59 patients. The 59 patients’ demographics and clinical characteristics.

|  | PMet (N=29) | PNoMet (N=30) | Total (N=59) | p value |
| --- | --- | --- | --- | --- |
| **Age at diagnosis** |  |  |  | 0.035<sup>1</sup> |
| Mean (SD) | 72.5 (10.9) | 78.0 (10.6) | 75.3 (11.0) |  |
| Median | 72.2 | 81.4 | 77.3 |  |
| Q1, Q3 | 66.7, 80.1 | 73.6, 83.7 | 67.7, 83.0 |  |
| Range | 44.8 - 90.8 | 52.6 - 93.6 | 44.8 - 93.6 |  |
| **Sex** |  |  |  | 0.761<sup>2</sup> |
| Female | 7 (24.1%) | 6 (20.0%) | 13 (22.0%) |  |
| Male | 22 (75.9%) | 24 (80.0%) | 46 (78.0%) |  |
| **Race** |  |  |  |  |
| N-Miss | 1 | 0 | 1 |  |
| White | 28 (100.0%) | 30 (100.0%) | 58 (100.0%) |  |
| **Immunosuppression** |  |  |  | 0.565<sup>2</sup> |
| N-Miss | 0 | 1 | 1 |  |
| Yes | 10 (34.5%) | 7 (24.1%) | 17 (29.3%) |  |
| No | 19 (65.5%) | 22 (75.9%) | 41 (70.7%) |  |
| **Tumor stage** |  |  |  | 0.604<sup>2</sup> |
| T2a | 16 (55.2%) | 15 (50.0%) | 31 (52.5%) |  |
| T2b | 12 (41.4%) | 15 (50.0%) | 27 (45.8%) |  |
| T3 | 1 (3.4%) | 0 (0.0%) | 1 (1.7%) |  |

1. Kruskal-Wallis rank sum test
2. Fisher's exact test

### Whole exome sequencing

Whole exome sequencing was performed on 61 tumors (PMet=29, PNoMet=32) and normal control tissues from 59 patients. We generated on average 119 ± 19 (mean±SD) million reads per sample with an average mapping rate of 99.8% ± 0.001% (mean±SD). The median depth of the sequencing is 155 (49-224), with 92% bases covered at 10X and 69% at 40X, on average. We identified 104,633 mutations within the coding regions across 17,094 genes, and 2,547 somatic variants were identified across 3 mutational callers with p-value <0.05 (see Methods, Supplemental Table 3 & Supplemental Figure 2). C>T transitions accounted for 86% of all SNV mutations (Supplemental Figure 2). Somatic mutations in the top 20 genes by Oncoplot were seen in 59 out of 61 tumor samples (96.72%) (Figure 1A). The most frequently mutated genes are seen in Figure 1A and include genes of interest: *TP53* (66%), *NOTCH1* (69%), and *FAT3* (66%). Gene function enrichment analysis (GFEA) found these mutated genes to be highly enriched in pathways in cell adhesion, endocytosis, regulation of small GTPase mediated signal transduction, and collagen fibril organization and extracellular matrix organization (Figure 1B, Supplemental Table 4). Driver mutation analysis was performed utilizing three mutational callers [MutSigCV (v1.41), OncodriveFML (v2.4.0), and OncodriveCLUSTL (v1.1.1)]. For integrative analysis, we prioritized 220 driver genes called from at least two mutational callers (Supplemental Table 5), including *TP53*, *CDKN2A*, *NOTCH1*, *SHC4*, *MIIP*, *CNOT1*, *C17orf66*, *LPHN22*, and *TTC16* as identified through all three calling systems (Figure 1C). Driver mutations common to both metastatic and non-metastatic cSCC include *TP53*, *NOTCH1*, *CDKN2A*, *MROH2A*, *KMT2D*, *PARD3*, and *FAT1* (Figure 1D). 220 Driver genes were found to be most enriched in pathways of replicative senescence, cellular response to UV, and cell adhesion and cell cycle (Figure 1E, Supplemental Table 6).

**Figure 1.**
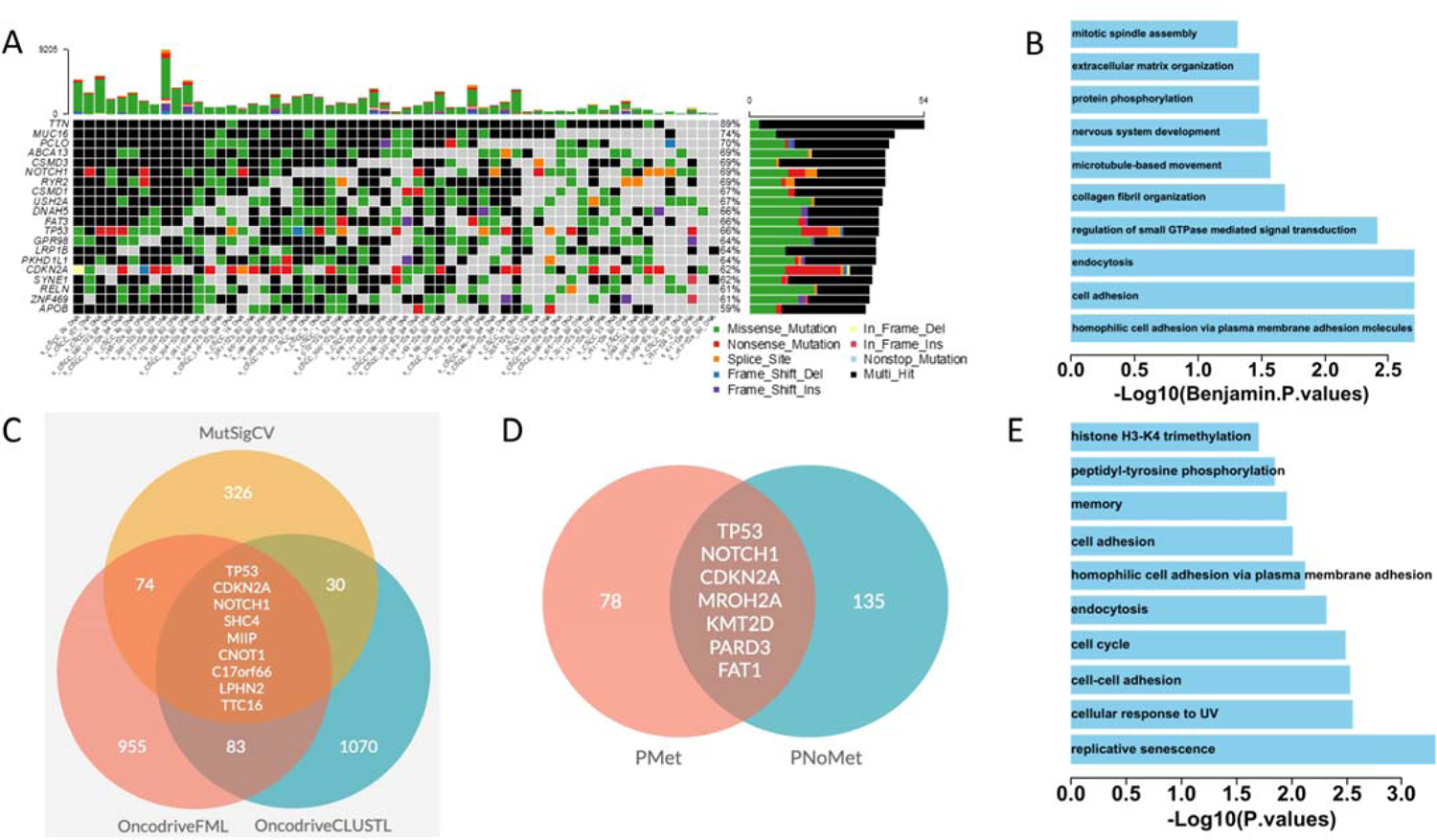
Exome-seq mutation analysis. A. Top 20 most common mutated genes in SCC patients. B. The enriched gene functions and pathways in 2547 mutated genes (potential drivers identified by at least 1 of 3 analysis tools) in the SCC patients (n=61). C. Prioritized mutated genes using 3 analysis tools. D. Mutated driver genes have been identified by a minimum of two out of three analysis tools in metastasis (PMet) and non-metastasis (PNoMet) patients. E. Gene functions and pathways enriched in the 220 mutated genes in Panel D.

### Whole transcriptome sequencing

Whole transcriptome sequencing was performed on a total of 53 primary tumors (PMet = 22, PNoMet = 31) and 22 normal controls from 50 patients (3 patients had multiple primary tumors) (Supplemental Figure 3). We generated on average 53 ± 27 (mean±SD) million reads per sample with an average mapping rate of 96% ± 2.37% (mean±SD). The total number of genes with non-zero counts was 44,307. We identified a total of 4,534 DEGs (1,966 up-regulated and 2,568 down-regulated) between tumor vs. controls |log2FC>1|, p-adj<0.05 (Supplemental Table 7). We identified a total of 39 DEGs (33 up-regulated and 6 down-regulated) when comparing primary tumors that metastasized vs those that did not metastasize (n=22 and n=31, respectively Supplemental Figure 4). Notable up-regulated genes in tumors vs. controls include: *MMP1* (Log2FC-5.6), *LAMC2* (Log2FC-3.8), 2, *CDKN2A* (Log2FC-3.6), and *MMP10* (Log2FC-7.8) and down-regulated genes include: *CFD* (Log2FC-3.3), *AR* (Log2FC-3.0), and *ABCA10* (Log2FC-2.4) (Figure 2A). GFEA showed up-regulation of immune and viral response, adaptive immune response, and cell division pathways (Figure 2B, Supplemental Table 8) and down-regulation of keratinization and epidermis development, intermediate filament organization, and metabolic processes pathways (Figure 2C, Supplemental Table 9). Master regulatory hub genes were identified amongst the top enriched pathways (Supplemental Figure 5). The top 50 regulatory hub genes are shown in Figures 2D and 2E, respectively. The top 50 gene sets are enriched for highly interacting genes in immune function.

**Figure 2.**
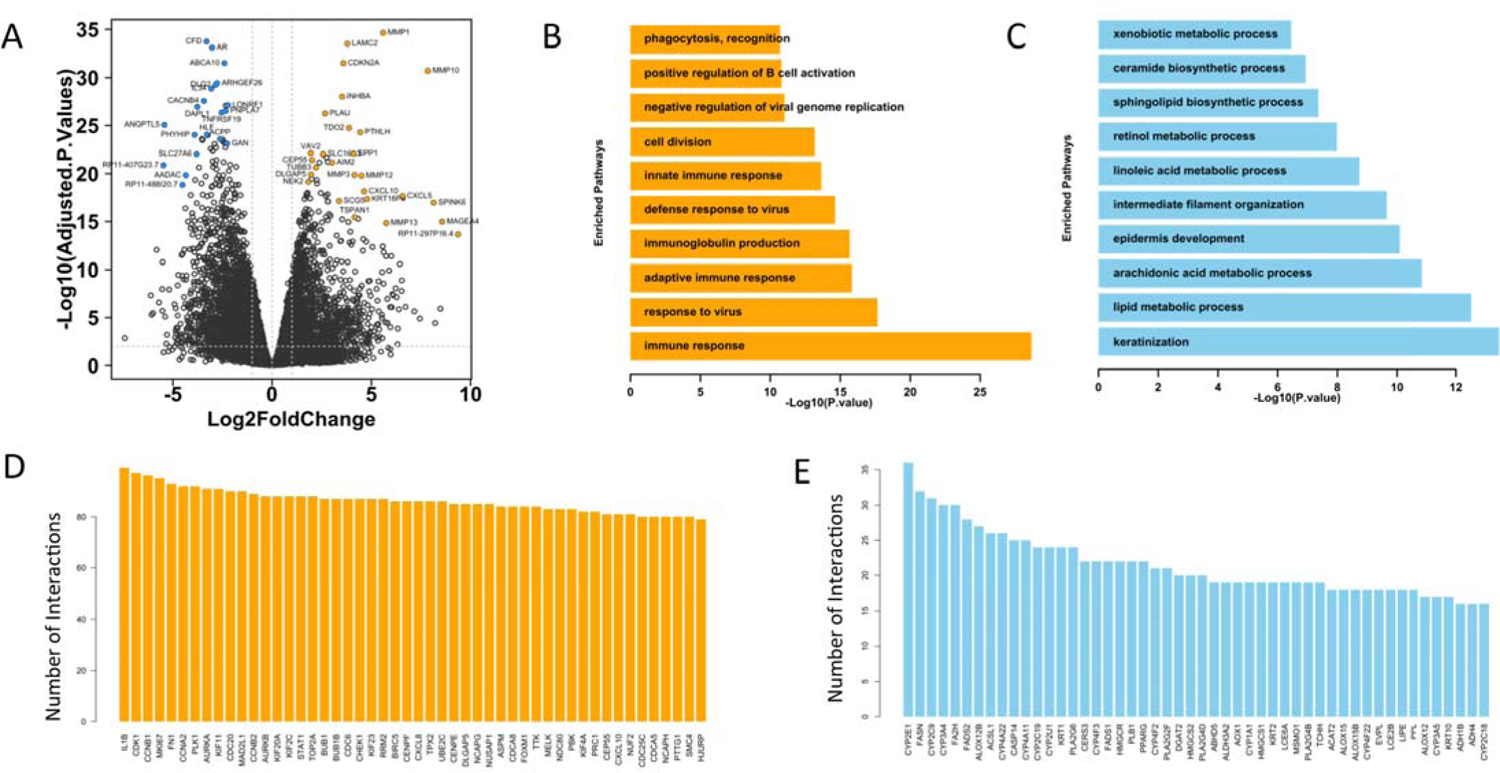
RNA-seq analysis. A. Volcano plot of differential genes of cSCC (n=53) vs. Controls (n=22) with highly significant genes highlighted. B. Top 10 enriched pathways in the upregulated differentially expressed genes (DEGs). C. Top 10 enriched pathways in the downregulated DEGs. D. Top 50 upregulated hub genes identified by gene interaction network analysis on genes in enriched upregulated pathways with Benjamin p value < 0.05. E. Top 50 downregulated hub genes in gene interaction network of genes in enriched downregulated pathways with Benjamin p value < 0.05.

### Integrative Analysis of Exome and Transcriptome Sequencing

A total of 183 critical genes were identified through integrative analysis of driver mutations, functional enrichment of mutated and differentially expressed genes, and master regulatory genes (See Methods, Figure 3A,B,& D, Supplemental Table 10). Through our combinatorial technique, we selected key driver genes to emphasize oncogenes, and crucial hub genes to feature those related to immune function.

**Figure 3.**
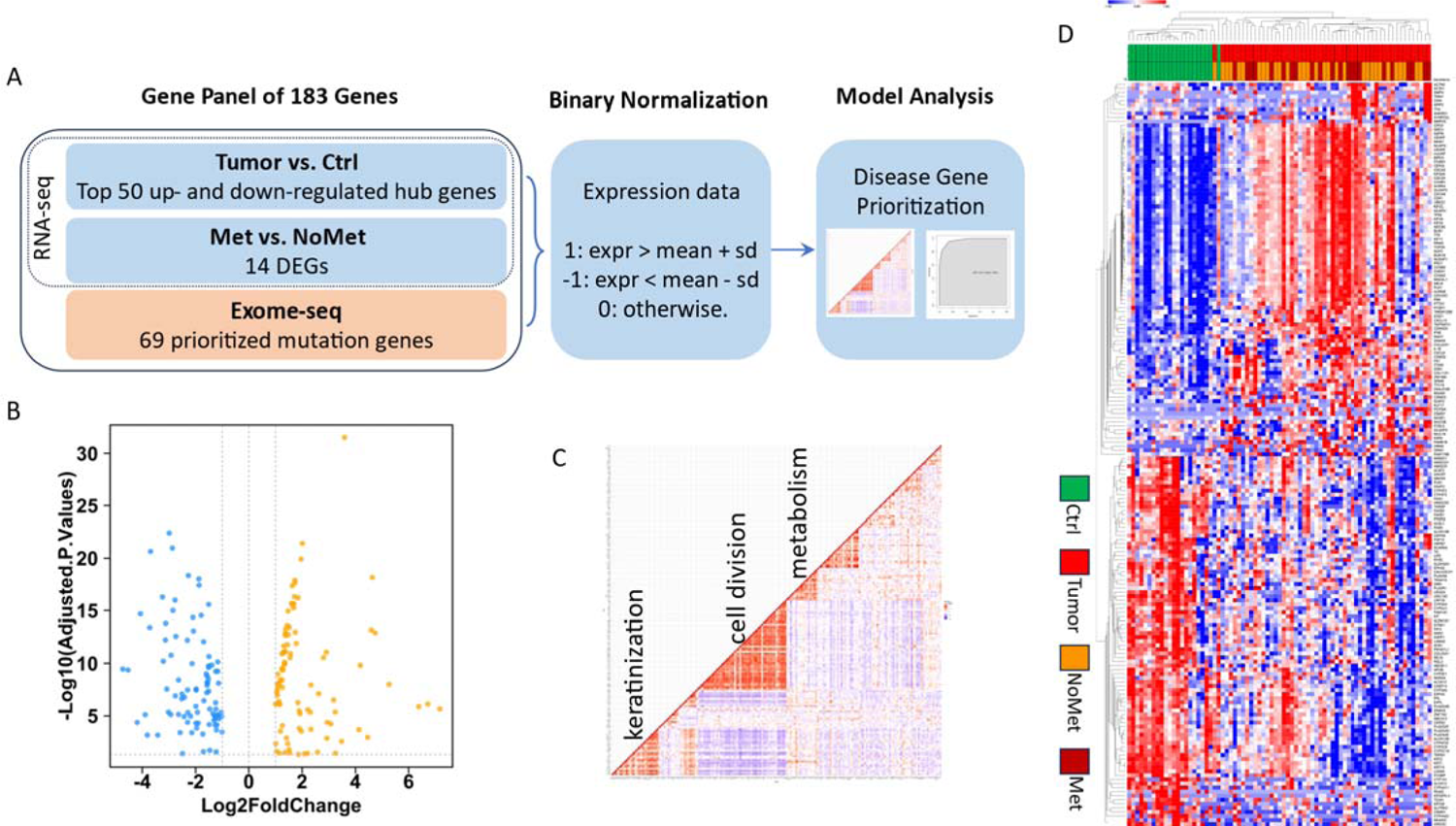
The establishment of SCC biomarker panel. A. Data integration pipeline for building SCC-specific marker panel of 183 genes for modeling analysis. The panel includes top 50 upregulated hub genes, top 50 downregulated hubs, top 14 DEGs between Met vs. NoMet from RNA-seq analysis, and 69 prioritized mutated genes identified by exome-seq analysis. B. Volcano plot show these 183 genes in RNA-seq analysis between SCC and control samples. C. The correlation heatmap of these 183 genes with main gene function highlighted for the 3 major clusters. D. Gene expression heatmap highlights the different patterns in SCC vs. Ctrl and Met vs. NoMet

### Derivation of gene panel

Using the PAM method, 183 genes clustered into 9 groups/clusters including 3 functionally enriched clusters in keratinization, cell division, and metabolism (Figure 3C, Figure 4, Supplemental Table 11). Fifty patients (22 Met, 28 NoMet; Supplemental Table 1, Supplemental Table 2) were used to model outcomes, and 16 genes were predictive of metastasis (Risk score ≥ 9 Met & Risk score < 9 NoMet). The Risk score for metastasis has an AUC of 97.1% (95% CI: 93.5% - 100%), sensitivity 95.5%, specificity 85.7%, and overall accuracy 90% (Figure 4A). Genes predictive of metastasis include: up-regulated: *CSAG1, UBE2C, ANKRD1, CSMD3, TOP2A, TPX2, KSR2, CSMD2, MMP20, PCSK1, TMEM150B*; up-regulated or no change: *RGL3, PLA2G6*; down-regulated: *ACSL1, TSGA10*, and *KIT* (Figure 4C,D; Supplemental Table 12). Univariate analysis with overall survival was performed using log-rank test for the 12 genes that were chosen by the model (Figure 4C,D; Supplemental Table 12). Eleven genes were chosen to generate the risk score for OS (Figure 4B). The Harrell’s C-statistic to predict OS is 80.8%. With each risk score increase, the risk of death increases by 2.47 (HR: 2.47, 95% CI: 1.64-3.74; p<0.001) after adjusting for age, immunosuppressant use, and metastasis status (Supplemental Table 13).

**Figure 4.**
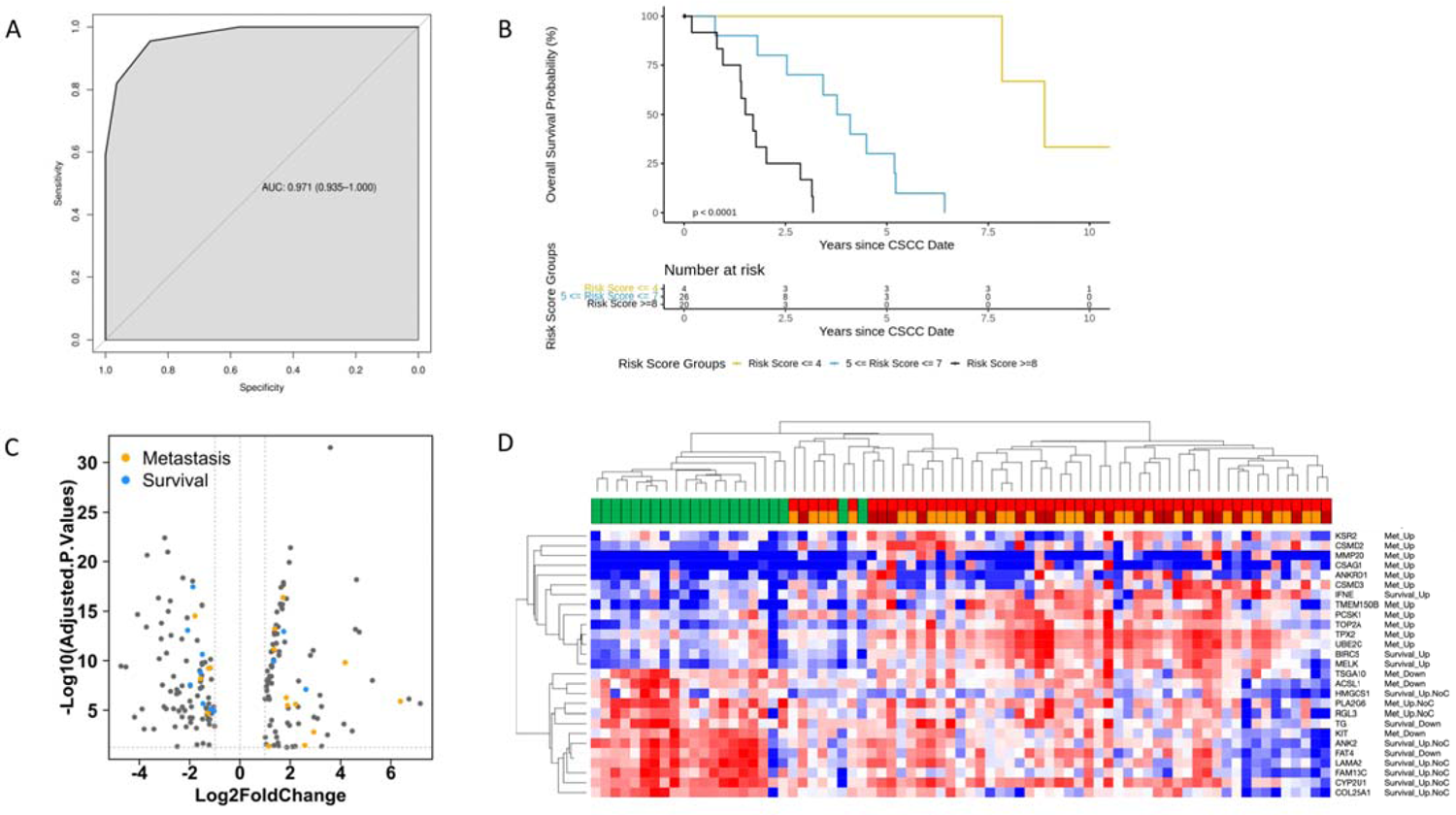
Modeling analysis of SCC biomarker panel. A. The AUC using the risk score to predict metastasis. B. The overall survival probability curve C. Modeling-selected genes related to metastasis and overall survival are highlighted in the volcano plot of 183 genes. D. The heatmap shows the expression patterns of these 27 genes selected by modeling analysis.

## Discussion

We have performed the first exome and transcriptomic sequencing on stage-matched, outcome-differentiated samples of intermediate and high-risk (T2a and T2b) cSCC. Through WES, we characterize the genomic landscape of intermediate to high-risk cSCC. In addition to commonly known driver mutations (*TP53*, *NOTCH1*, *CDKN2A*, *FAT1*), we identified unique driver genes including *MROH2A*, *KMT2D*, *PARD3*, *SHC4*, *MIIP*, *CNOT1*, *C17orf66*, *LPHN2*, and *TTC16* [11, 34–36]. Some of these unique driver genes were found to be driver genes in lung SCC (*KMT2D, LPHN2*), bladder cancer (*PARD3*), colorectal cancer *(MIIP*), and hypothesized to be a potential driver in cSCC (*PARD3*) [14, 37–40]. Namely, *KMT2D,* which codes for a histone methyltransferase, has been implicated in several different types of cancers and more recently has been suggested to be driving lung squamous cell carcinoma through epigenetic modifications [38, 41]. Others that were found to be implicated in carcinogenesis include *MROH2A*, *SHC4*, *CNOT1*, and *C17orf66*, but further investigation is needed to define their roles [42–45]. In total, 220 driver genes were found to be enriched in replicative senescence, cellular response to UV, cell adhesion, and cell cycle pathways. We found that UV mutations accounted for most of the somatic mutations. Notably, when we performed driver enrichment analysis, UV-driven processes were found to be a primary enriched pathway that drove the disease. Mutation load has been correlated with cumulative UV exposure as well as keratinocyte cancer burden [34]. Our study is the first to perform driver enrichment analysis that establishes the critical role of UV in driving aggressive cSCC. We found other well-known cancer pathways when performing driver gene enrichment analysis such as replicative senescence, cell adhesion, and cell cycle pathways [11, 34–36].

Through whole transcriptome sequencing, we identified DEGs between tumor vs. control and between primary tumors that metastasized vs those that did not metastasize, many of which have not been previously reported in cSCC. Significantly upregulated DEGs included genes in the matrix metalloproteinase (*MMP*) family such as *MMP1*, *MMP3*, *MMP10*, *MMP12*, and *MMP13*, highlighting the importance of extracellular matrix remodeling and stromal response in carcinogenesis [46]. We then leveraged interactional gene networks to identify master regulatory hub genes and discovered functionally relevant pathways in cSCC development, including immune- and stromal-related genes. Consistent with previous studies, immune response, keratinization, and metabolic pathways were among the most highly enriched [35].

We implemented a comprehensive approach by selecting key driver genes and hub genes, highlighting oncogenes and genes critical to immune function, respectively. We identified 183 genes critical to cSCC carcinogenesis that were predictive of metastasis and survival [11–15, 47–54]. We found three distinct functional clusters of genes involved in metabolism, keratinization, and cell division. Genes involved in metabolic pathways are known to be down-regulated in metastatic cSCC, we are the first to show they predict overall survival [15]. We performed modeling and identified 16 genes predictive of metastasis with an accuracy of 90%, specificity of 86%, and sensitivity of 96%. We also identified 11 genes that are independently associated with lower survival with a survival prediction accuracy of 81%. The predictive genes were up-regulated in common pathways by GFEA including: cell division (*BIRC5*, *UBE2C*, *TPX2*), apoptotic process (*BIRC5*, *MELK*, *TPX2*), and mitotic cell cycle (*BIRC5*, *TPX2*). Unique genes were up-regulated in immune response (*IFNE*), cellular response to hypoxia (*ANKRD1*), cell-cell signaling (*PCSK1*), and cell proliferation pathways (*MELK*). The genes were downregulated in common pathways by GFEA including: lipid metabolic process (*HMGCS1*, *PLA2G6*), response to drug (*HMGCS1*, *ACSL1*), signal transduction (*ANK2*, *KIT*), as well as pathways of interest such as cell-cell adhesion (*FAT4*), cell adhesion (*LAMA2*), fibroblast growth factor receptor signaling pathway (*FAT4*).

Previous work has identified GEP signatures that may predict metastatic risk, but these have not been able to predict both metastasis and survival [17, 18, 55]. Other GEP derivations were inclusive of large numbers of low-risk tumors unlikely to metastasize, only controlling for age, sex, and tumor location, neglecting to control for stage [18]. Intermediate to high-risk tumors (BWH T2a & T2b) are known to have more outcome variability, making prediction and risk-stratification increasingly difficult [5]. Our primary tumors were phenotypically similar but were stage- and immunosuppression-matched to identify the molecular and genetic differences underlying their behavior and outcomes. We identified the enrichment of unique, critical pathways of cSCC carcinogenesis that predict outcomes of stage-matched, outcome differentiated tumors. Our GEP profile has the potential to guide management through the accurate identification of at-risk tumors as well as the ability to predict survival in individuals with intermediate to high-risk cSCC. Future studies examining the performance of our assay across the spectrum of cSCC is warranted.

## Data Availability

All data produced in the present study are available upon reasonable request to the authors.

## Supplemental Figures

**Supplemental Figure 1.**
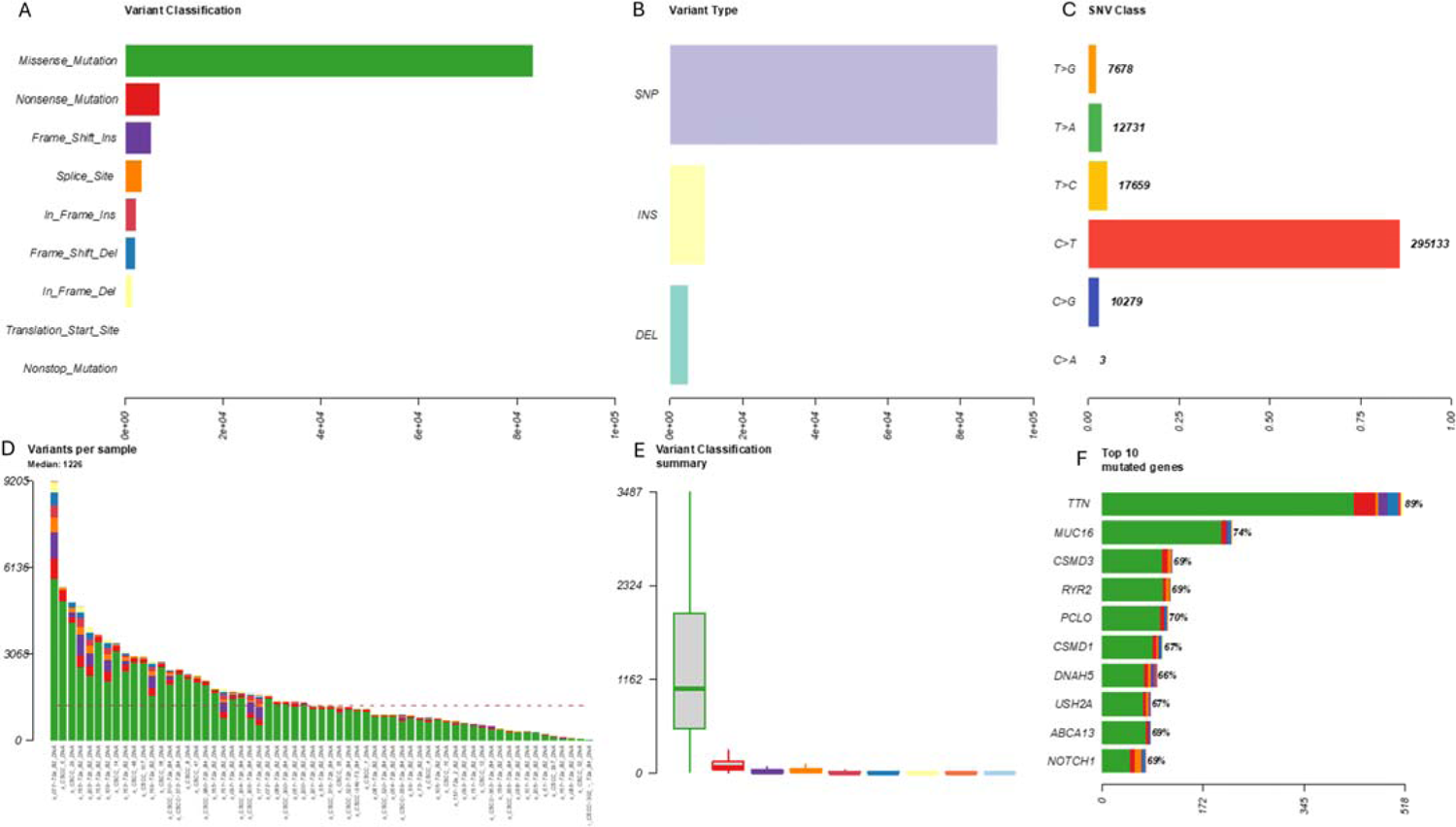
Summary of the somatic variants identified from 61 tumor samples A. Barplot showing the classification of all somatic variants in coding regions B. Barplot showing number of somatic variants in each type C. Barplot showing number of SNV variants in six different conversions (Transitions and Transversions) D. Barplot showing number of somatic variants in each tumor samples E. Boxplot showing the distribution of the somatic mutations in each category among tumor samples (colors corresponding to each category in A) F. Stacked barplot showing fraction of the somatic mutation in each category for top 10 mutated genes (colors corresponding to each category in A)

**Supplemental Figure 2.**
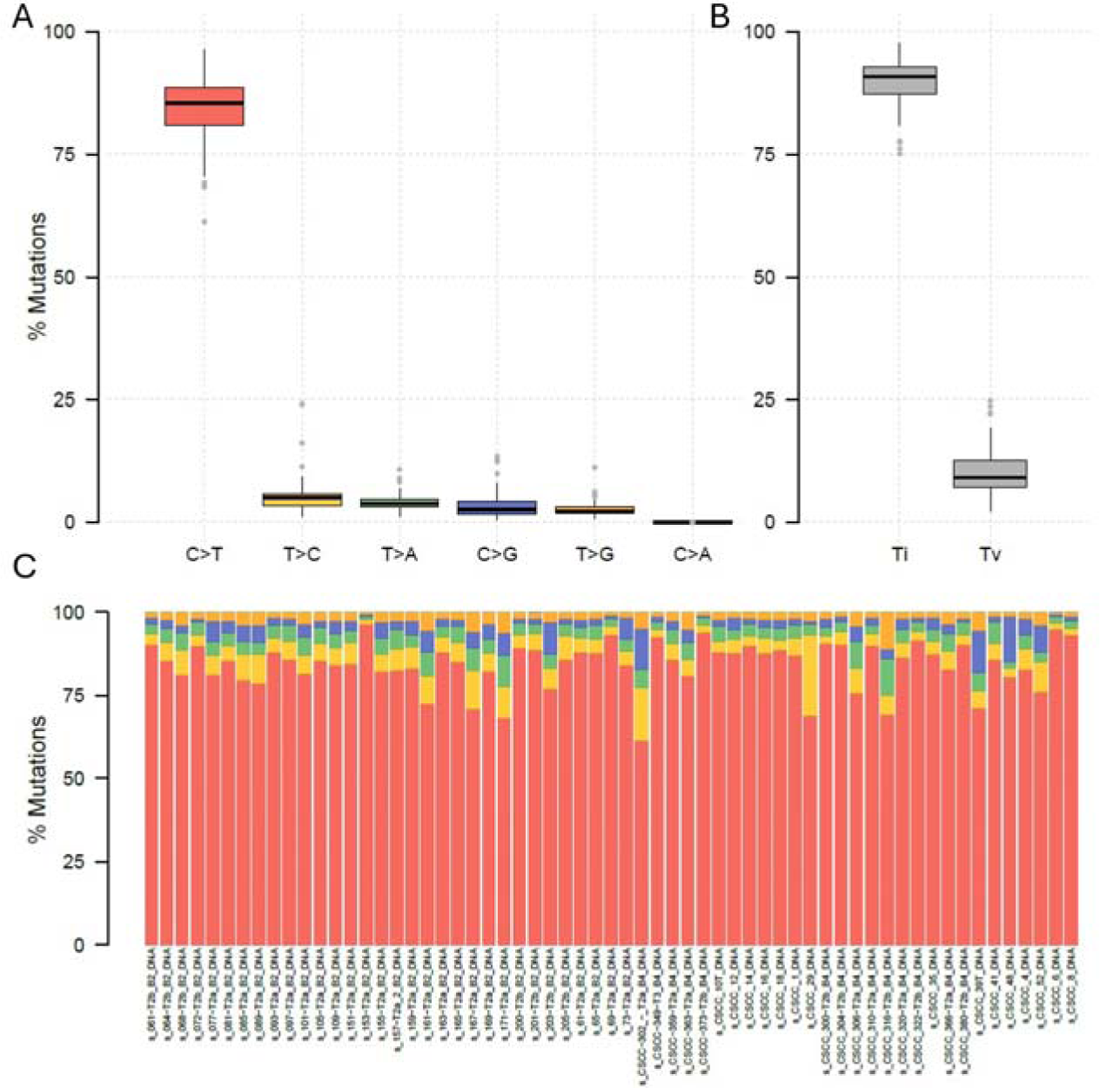
Transitions and Transversions summary of all SNV mutations across 61 tumor samples A. Overall distribution of six different conversions B. Boxplot showing overall Transition and Transversion. C. Stacked barplot showing fraction of conversion in each of the tumor samples (colors corresponding to six different conversions in A)

**Supplemental Figure 3.**
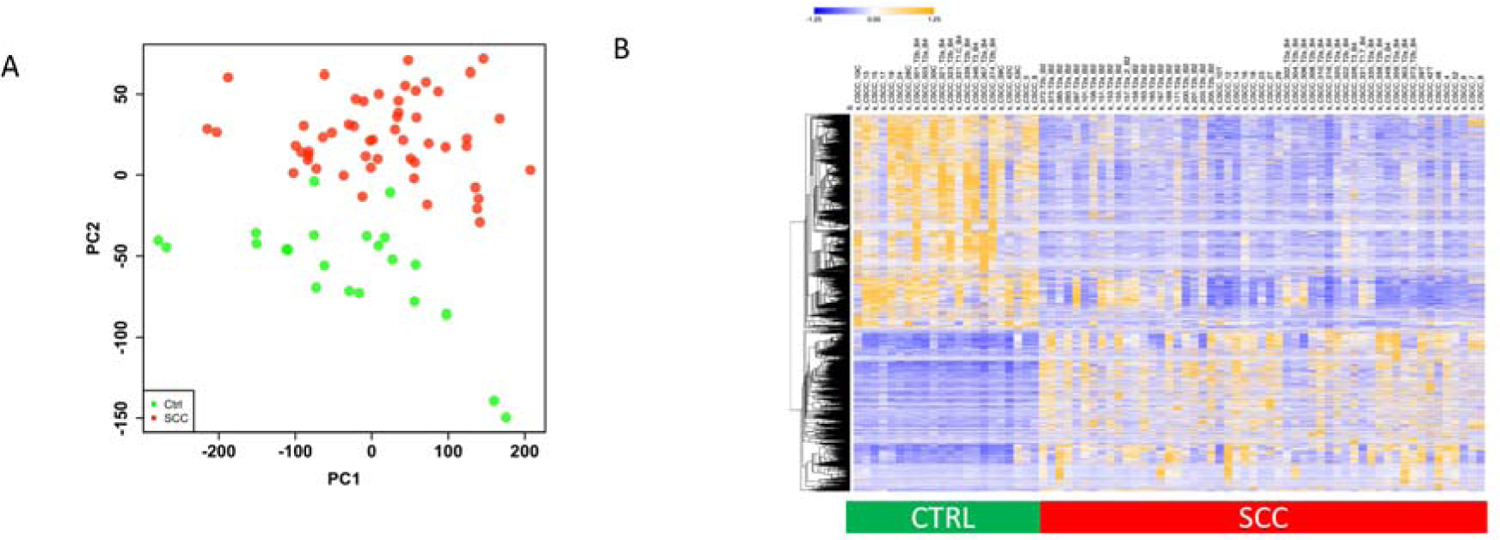
Transcriptomic profiling using RNA-seq in SCC patient. A. PCA analysis of all 75 sample. B. Heatmap of DEGs between tumor and control. B. Differentially expressed genes between SCC patient skin tissue samples (SCC, n=53) and healthy control skins (CTRL, n=22)

**Supplemental Figure 4.**
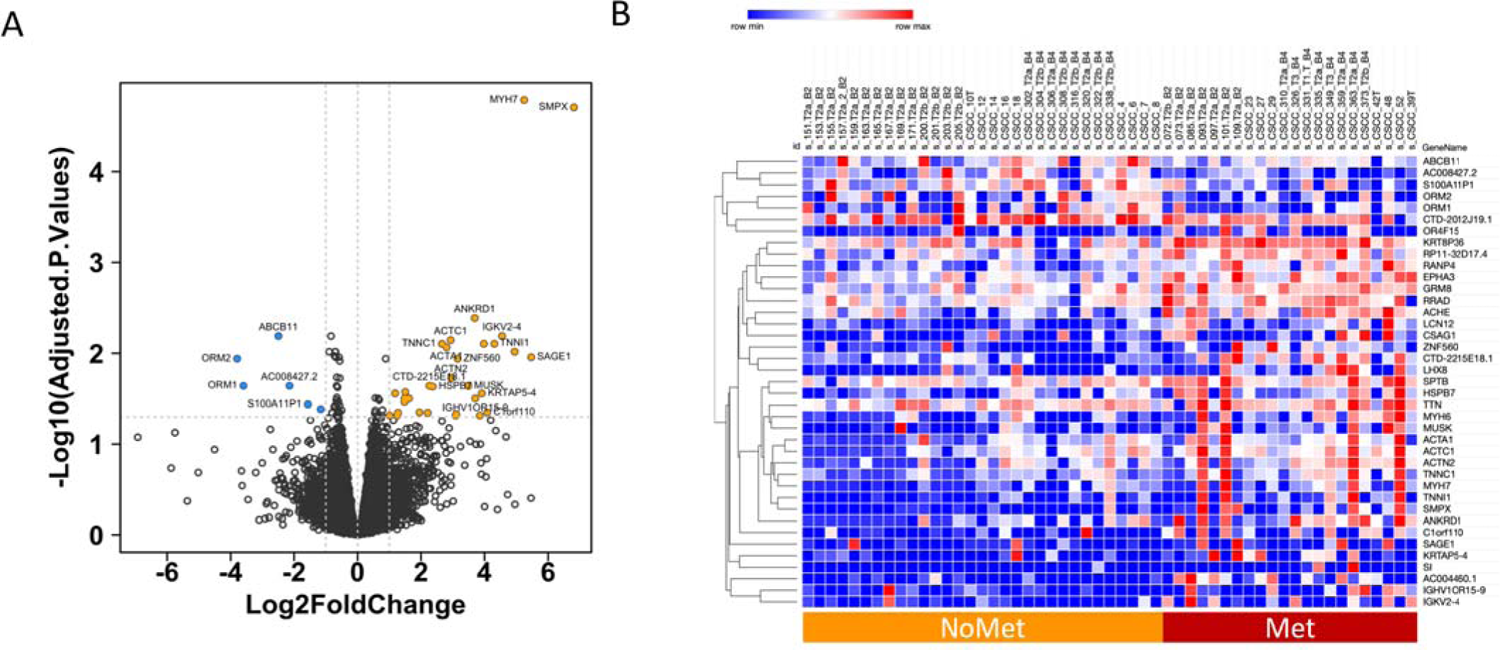
Transcriptomic profile comparison between metastatic and non-metastatic tumors. A. Volcano plot of differential analysis of Met (n=22) vs. NoMet(n=31). B. Top Differential Genes between Met and NoMet, with adj.p < 0.05 & |Log2FC|>1

**Supplemental Figure 5.**
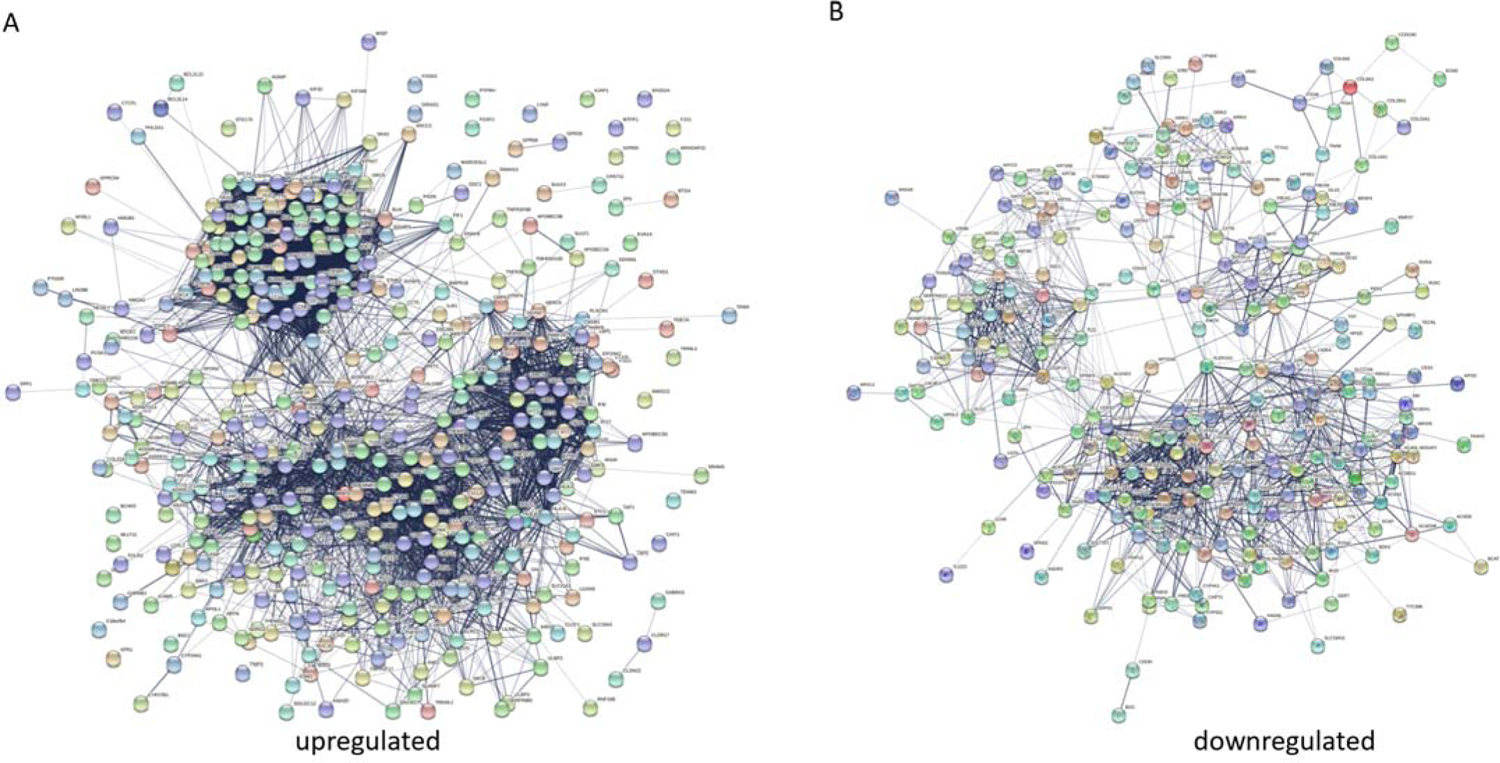
Gene interaction network of genes in top enriched pathways. A. Gene-gene interaction networks in the genes of top upregulated enriched pathways with Benjamin p value < 0.05 B. Gene-gene interaction networks in the genes of top downregulated enriched pathways with Benjamin p value < 0.05

## Supplemental Tables

**Supplemental Table 1:** Patient demographics-The 53 RNA samples are from 50 patients. The 50 patients’ demographics and clinical characteristics.

|  | PMet (N=22) | PNoMet (N=28) | Total (N=50) | p value |
| --- | --- | --- | --- | --- |
| **Age at CSCC** |  |  |  | 0.165<sup>1</sup> |
| Mean (SD) | 73.4 (12.4) | 78.1 (9.2) | 76.0 (10.9) |  |
| Median | 75.3 | 79.8 | 77.9 |  |
| Q1, Q3 | 67.8, 82.4 | 73.7, 83.8 | 71.3, 83.2 |  |
| Range | 44.8 - 90.7 | 58.2 - 91.6 | 44.8 - 91.6 |  |
| **Sex** |  |  |  | 0.197<sup>2</sup> |
| Female | 8 (36.4%) | 5 (17.9%) | 13 (26.0%) |  |
| Male | 14 (63.6%) | 23 (82.1%) | 37 (74.0%) |  |
| **Race** |  |  |  |  |
| White | 22 (100.0%) | 28 (100.0%) | 50 (100.0%) |  |
| **Immunosuppressant Use** |  |  |  | 1.000<sup>2</sup> |
| Yes | 7 (31.8%) | 8 (28.6%) | 15 (30.0%) |  |
| No | 15 (68.2%) | 20 (71.4%) | 35 (70.0%) |  |
| **Disease stage** |  |  |  | 0.232<sup>2</sup> |
| T1 | 1 (4.5%) | 0 (0.0%) | 1 (2.0%) |  |
| T2a | 10 (45.5%) | 13 (46.4%) | 23 (46.0%) |  |
| T2b | 9 (40.9%) | 15 (53.6%) | 24 (48.0%) |  |
| T3 | 2 (9.1%) | 0 (0.0%) | 2 (4.0%) |  |

<sup>1.</sup> Kruskal-Wallis rank sum test
<sup>2.</sup> Fisher's exact test

Supplemental Table 2: Sample Annotation in 75 RNA-seq Samples in cSCC and Control Samples

Supplemental Table 3: Somatic Variant List Identified with Exome-seq

Supplemental Table 4: Enriched Pathways in Somatic Mutations

Supplemental Table 5: 220 Driver Genes Identified by 2 Calling Systems

Supplemental Table 6: Enriched Pathways in 220 Driver Mutations

Supplemental Table 7: DEGs in SCC vs. Ctrl

Supplemental Table 8: Enriched Pathways in Up-regulated DEGs in cSCC vs. Control

Supplemental Table 9: Enriched Pathways in Down-regulated DEGs in cSCC vs. Control

Supplemental Table 10: 183 Genes Identified by Integrative Analysis

Supplemental Table 11: Enriched Pathways in Clusters in Correlation Heatmap of 183 Gene Panel

Supplemental Table 12: Twenty-seven Genes Predictive of Outcome from 183 Gene Panel

**Supplemental Table 13:** Overall Survival Multivariable Analysis.

| Characteristic | HR<sup>1</sup> | 95% CI<sup>1</sup> | p-value |
| --- | --- | --- | --- |
| Risk Score based on 11 genes (every unit increased) | 2.47 | 1.64, 3.74 | <0.001 |
| Age at CSCC | 1 | 0.96, 1.04 | 0.8 |
| Immunosuppressant Yes vs. No | 1.36 | 0.43, 4.28 | 0.6 |
| Metastasis Status Yes vs. No | 2.06 | 0.80, 5.31 | 0.14 |
| <sup>1</sup> HR = Hazard Ratio, CI = Confidence Interval |  |  |  |

